# Enhancing Title and Abstract Priority Screening Through EdRoSi AI Pipeline

**DOI:** 10.64898/2026.06.26.26356718

**Authors:** Pacôme Lecot, Hélène Tonoli-Catez, Arthur Noseda, Thibault Buisse, Stéphanie Chanut, Isabelle Chanel

## Abstract

The exhaustive identification of evidence is central to systematic reviews, but the screening of titles and abstracts remains particularly labor intensive. Priority screening, an active learning approach that ranks records by estimated relevance, has emerged as an effective strategy to reduce screening workload. Its efficiency is commonly quantified using work saved over sampling at 100% recall (WSS@100%), representing the percentage reduction in effort compared with random screening. Although modern priority-screening models achieve high efficiency on many benchmark datasets, some reviews still exhibit low WSS@100%, indicating suboptimal retrieval. Our study sought to improve the retrieval of all relevant articles in challenging datasets to ensure better generalization of priority screening. We first showed using SYNERGY benchmark datasets that while the most advanced ELAS_h3 priority screening model from state-of-the-art ASReview LAB v.2 open-source software, efficiently retrieved most relevant articles, it struggled with the rare, final ones in challenging datasets. To address this, we tested a hybrid approach entitled EdRoSi AI: using ELAS_h3 for early retrieval and then applying supervised fine-tuning to the biomedical transformer BioMed-RoBERTa-base with these relevant articles to enhance the detection of the remaining difficult cases. We found that fine-tuning BioMed-RoBERTa-base model with 10 late-identified relevant and 10 hard irrelevant study titles and abstracts, enabled faster retrieval of articles of interest compared to ELAS_h3 alone. This approach increased WSS@100% from 46.5% (SD 0.0%) to 83.3% (SD 0.4%), while adding only an average of 22 minutes of computational time for fine-tuning and inference. The EdRoSi AI priority screening pipeline could be valuable for situations requiring highest possible recall. It could be particularly useful in scoping reviews with broad or diverse topics where traditional priority screening methods may miss subtle relevance signals. Further work should define a data-driven stopping rule for ending screening once the fine-tuned domain-specific transformer is applied at the final stage and assess generalizability across additional challenging datasets.

## Introduction

Guidelines in medicine are constructed from high-quality, up-to-date evidence synthesized through systematic reviews^1,2^. One particularly resource-intensive stage of systematic review development is the screening of titles and abstracts from potentially relevant articles, a process that can require over 48 hours of work per expert for the screening of 10 000 articles^3^.

To address this bottleneck, several approaches have been developed to semi-automate or fully automate the title and abstract screening stage^4^. Most binary text classification methods rely on supervised machine learning (ML) using either traditional models or deep learning architectures^4–6^. These methods have demonstrated the ability to achieve over 99% recall in some datasets, meeting the minimum standards set by the highly trusted Cochrane organization^5^. Despite their effectiveness, these models often require large, domain-specific training corpora, which limit their generalizability across diverse topics. Alternatively, generative artificial intelligence (AI) models, which utilize self-supervised learning, have recently emerged as promising tools for title and abstract screening^7–9^. Nevertheless, according to the Responsible AI in Evidence SynthEsis (RAISE) principles, they do not yet seem to be ready for large adoption due to performance variability linked to prompt quality and methodological complexity; the lack of human oversight and the lack of transparency which could compromise result reproducibility^10–12^.

Priority screening, an active learning-based approach, remains a trusted method for accelerating the title and abstract screening process and is endorsed by PRISMA guidelines^13^. In this iterative process, users begin with a minimal set of relevant and irrelevant studies, which are used to train the model^14–16^. The model then sequentially recommends the most likely relevant articles, retraining after each decision until all relevant items are identified. This strategy is integrated into various screening platforms, such as Abstrackr, Colandr, Rayyan, RobotAnalyst, Research Screener, DistillerSR, and RobotReviewer. More recently, transparent open-source softwares, like ASReview and DenseReviewer have been developed specifically for priority screening^15,17,18^.

Studies have shown that priority screening can significantly reduce screening time compared to screening articles in a random order^15,19^. ASReview can significantly reduce the workload required to achieve full recall of relevant articles, by as much as 93% in certain datasets, according to the Work Saved over Sampling at 100% recall (WSS@100%) metric. Yet its efficiency drops on benchmark datasets like those from *Cohen* and *Kwok* datasets, where WSS@100% falls to 38% and 42%, respectively^15^. Developing new priority screening pipelines to retrieve more efficiently all relevant articles in these challenging datasets is important. Indeed, it would broaden the applicability of priority screening, especially for systematic reviews with “hard-to-find” relevant articles.

In our study, we showed that a pre-trained transformer model, fine-tuned with only 10 relevant and 10 irrelevant biomedical articles previously identified using static text-representation active learning methods, was sufficient to greatly improve the retrieval of “hardest-to-find” studies during priority screening compared to state-of-the-art active learning models.

## Materials and Methods

### Validation Datasets

Simulations were conducted using four publicly available validation datasets from the SYNERGY collection selected to represent contrasting screening efficiency, as quantified by the Work Saved over Sampling at 100% recall (WSS@100%)^15,20^. Two datasets exhibited WSS@100% values inferior to 50% and were therefore considered *challenging*, whereas two datasets showed WSS@100% values superior to 90% and were considered *less challenging*.

#### Datasets with Low WSS@100% (“Challenging” Datasets)

The first challenging dataset was derived from *Cohen* dataset, corresponding to a systematic review on the efficacy of angiotensin-converting enzyme (ACE) inhibitors. This dataset originates from a curated collection of systematic review datasets in the medical sciences^14^. It consists of a static subset of 2,544 publications extracted from the TREC 2004 Genomics Track document corpus, which includes MEDLINE records published between 1994 and 2003. The use of a fixed corpus enables full replicability of results. A total of 41 publications were deemed relevant in the original review, corresponding to a prevalence of 1.61%.

The second challenging dataset was obtained from *Kwok* dataset and corresponds to a systematic review of viral metagenomic next-generation sequencing studies conducted in common livestock species, including cattle, small ruminants, poultry, and pigs^21^. Following deduplication, the initial search yielded 2,481 studies, of which 120 were classified as relevant, resulting in a prevalence of 4.84%.

#### Datasets with High WSS@100% (“Less Challenging” Datasets)

The third dataset was drawn from *Bos* dataset and relates to a systematic review and meta-analysis of prospective population-based studies investigating the associations of white matter hyperintensities, covert brain infarcts (*i.e.,* clinically silent infarcts), and cerebral microbleeds with the risk of all-cause dementia or Alzheimer’s disease^22^. After deduplication, the initial search comprised 5,553 studies, among which 10 were identified as relevant, corresponding to a prevalence of 0.18%.

The fourth dataset originated from *Van de Schoot* dataset^23,24^. This dataset represents a review of longitudinal studies applying unsupervised machine-learning techniques to longitudinal self-reported symptom data on post-traumatic stress following trauma exposure. The initial search retrieved 5,782 studies, of which 38 met the inclusion criteria, yielding a relevance prevalence of 0.66%.

### Assessment of ASReview LAB v.2 Screening Efficiency

In this study, we performed AI-assisted title and abstract screening using ASReview LAB v.2 through the ASReview Python API (*i.e*., the Application Programming Interface that enables programmatic control of the ASReview active learning workflow, including data ingestion, model configuration, iterative training, and ranking export)^17^. We evaluated two ELAS configurations available in ASReview: ELAS_u4, which represents an active learning pipeline combining a TF-IDF (term frequency–inverse document frequency) text feature extractor with a linear support vector machine (SVM) classifier, and ELAS_h3, which uses the *mxbai-embed-large-v1* transformer embedding model as the feature extractor coupled to an SVM classifier. For each configuration, ASReview iteratively updates the classifier as labels accumulate and continuously re-ranks the remaining unlabeled records, thereby prioritizing records predicted to be most relevant for screening.

Model performance was evaluated using simulation mode, which replays the screening process against. It enables computation of WSS@100%^15^. After ranking all records according to the model-generated relevance scores, the ranking was iteratively updated until all relevant studies were identified (100% recall). WSS@100% was then calculated as:

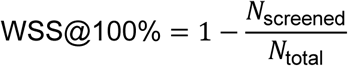

where *N*_screened_is the number of records that must be screened to retrieve all relevant studies, and *N*_total_is the total number of records in the dataset. Higher WSS@100% values indicate greater screening efficiency, reflecting the model’s ability to rank relevant studies early.

### Fine-Tuning of BioMed-RoBERTa-base Model

We conducted local fine-tuning of a transformer model pre-trained on biomedical literature (BioMed-RoBERTa-base) to rank records by their relevance. BioMed-RoBERTa-base developed by Allen Institute for Artificial Intelligence (allenai/biomed_roberta_base), was downloaded from Hugging Face online platform on October 7^th^, 2025^25^. The input text for each record was built by concatenating the title and abstract into a single sequence, after excluding rows with missing/empty title or abstract, ensuring that the model was trained only on informative textual content.

#### Data preparation and tokenization

All texts were tokenized using the RoBERTa tokenizer associated with the base model directory. Tokenization used truncation with a maximum sequence length of 512 tokens, which matches the model’s standard context window and allows capturing most biomedical abstracts while preventing out-of-memory errors. Dynamic padding was applied at batch time via a padding collator, minimizing unnecessary computation for shorter sequences.

#### Model and optimization procedure

We used AutoModelForSequenceClassification with two output labels (relevant/irrelevant). Fine-tuning was performed in a single-shot setting, corresponding to one training run without cross-validation, on balanced number of relevant and irrelevant articles. The use of a single-shot setting was warranted due to the limited number of labeled examples, as this approach optimizes the quantity of data available for parameter updates. The tokenizer was saved together with the fine-tuned model to preserve the exact text preprocessing and token-to-ID mapping required to reproduce training and ensure consistent inference when reloading the model artifact.

#### Rationale for hyperparameter choices for small, labeled fine-tuning datasets

Because our fine-tuning dataset contains a limited number of relevant and irrelevant examples, we selected conservative hyperparameters aimed at reducing overfitting and stabilizing optimization:

- **Low learning rate = 5×10⁻⁶.** With few labeled examples, a typical “standard” transformer fine-tuning learning rate (e.g., 2–5×10⁻⁵) can cause large parameter updates and rapid memorization of the training set. We therefore used a smaller learning rate to adapt the pretrained representations more gradually, preserving general biomedical language knowledge while allowing the classification head and upper layers to specialize to subtle relevance cues.
- **Small batch size = 4.** Small datasets are particularly sensitive to optimization dynamics and memory constraints with 512-token sequences. A batch size of 4 reduces memory usage and enables training on typical workstations while still providing sufficiently frequent gradient updates. Additionally, smaller batches introduce mild gradient noise that can act as a regularizer, which is beneficial when data are scarce.
- **Moderate number of epochs = 6.** With limited data, too few epochs may underfit, whereas too many epochs can overfit quickly. We used 6 epochs as a pragmatic compromise to allow the classifier to converge while keeping training duration modest.
- **Weight decay = 0.01.** We applied L2 regularization through weight decay to reduce model complexity and mitigate overfitting, which is a common risk in small-sample fine-tuning.

### Inference with Fine-Tuned BioMed-RoBERTa-base Model

For each citation, the model input was constructed by concatenating the title and abstract into a single text sequence, which was then tokenized using the tokenizer associated with the selected fine-tuned checkpoint. Tokenization was performed with truncation to a maximum length of 512 tokens and dynamic padding at batch time.

Inference was performed using a previously fine-tuned AutoModelForSequenceClassification (two output labels). Predictions were generated in evaluation mode using mini-batches, and probability that a given article belongs to the positive (relevant) class was derived by applying a softmax function to the model logits.

### Screening Efficiency Assessment of Hybrid Priority Screening Approach ASReview LAB v.2 Model Combined with Fine-Tuned BioMed-RoBERTa-Base

The performance of the hybrid priority screening approach was evaluated using Work Saved over Sampling at 100% recall (WSS@100%). In this setting, the total number of screened articles required to achieve complete recall corresponded to the sum of articles already screened during the ASReview ELAS_h3 workflow and the additional new articles screened following re-ranking by the fine-tuned BioMed-RoBERTa-base model.

## Results

### Evaluation of Priority Screening Efficiency for Each Relevant Articles Using ASReview LAB v.2 Models in Challenging and Less Challenging Datasets

The ASReview Python API was utilized to simulate systematic reviews, initiating each simulation with one randomly selected relevant article and one randomly selected irrelevant article. Three simulations were conducted for each benchmark dataset under consideration, and the mean WSS@100% as well as its standard deviation were calculated. The ELAS_h3 model demonstrated superior WSS@100% values across all validation datasets when compared to ELAS_u4 (Supplemental Table 1). The absolute mean increases in WSS@100% for ELAS_h3 over ELAS_u4 were 5.060% for *Bos*, 12.055% for *Van De Schoot*, 15.409% for *Cohen*, and 14.107% for *Kwok* datasets.

Despite higher WSS@100% with ELAS_h3 compared to ELAS_u4, it remained low for challenging datasets: *Cohen* dataset (mean 46.528%, SD 0.023%) and *Kwok* dataset (mean 45.089%, SD 0.047%) (Supplemental Table 1). In contrast, less challenging datasets showed much higher WSS@100% values: *Bos* dataset (mean 94.598%, SD 0.036%) and *Van De Schoot* dataset (mean 95.105%, SD 0.135%) (Supplemental Table 1). ELAS_h3 improved WSS@100% for challenging datasets, however, it did not enable to efficiently retrieve all relevant articles. Specifically, the amount of work saved over sampling is only half as much compared to less challenging datasets.

We next investigated how often “hard-to-find” relevant articles appeared in challenging datasets and where their identification was most difficult during screening. Screening efficiency was measured by counting how many irrelevant articles were reviewed before finding each relevant one.

Using ELAS_h3 and ELAS_u4, in *Cohen* and *Kwok* datasets, only a few irrelevant articles needed to be screened to find most relevant articles early in the process (Figures 1B and 1D). Nevertheless, the process of identifying the final relevant articles necessitated the review of a substantially larger number of irrelevant articles. Although ELAS_h3 demonstrated greater efficiency compared to ELAS_u4 in this context, the quantity of irrelevant articles remained substantial: on average, 967 (SD 2) were encountered prior to identifying the “hardest-to-find” relevant article in the *Cohen* dataset, and 254 (SD 5) in the *Kwok* dataset.

**Figure 1:**
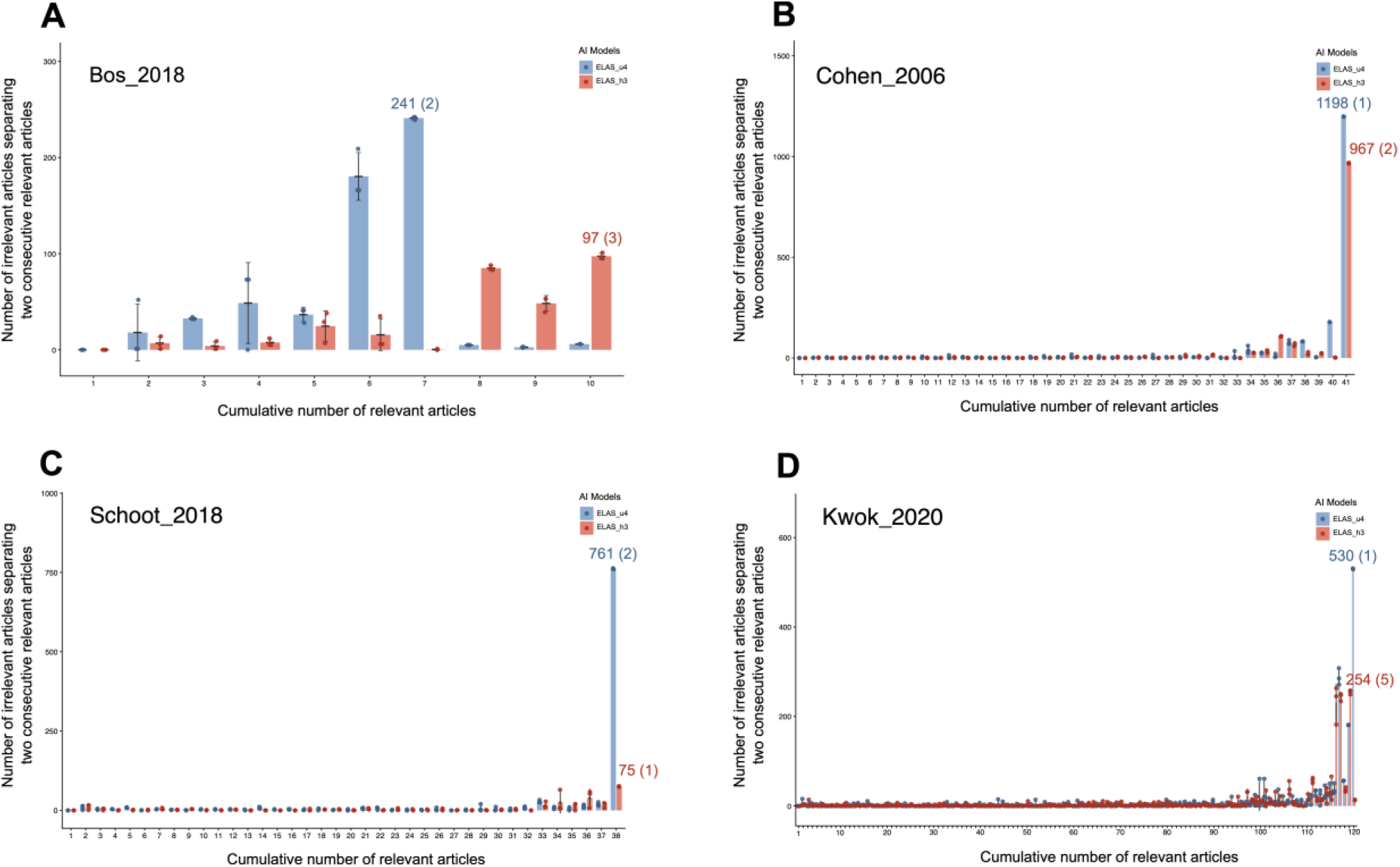
Evaluation of Priority Screening Efficiency for Each Relevant Articles Using ASReview LAB v.2 Models in Challenging and Less Challenging Datasets. Panels A–D showed screening efficiency, defined by the number of irrelevant articles separating two consecutive relevant articles (y–axis), plotted against the cumulative number of relevant articles identified (x–axis) for four benchmark datasets. In this context, a higher number of intervening irrelevant articles corresponded to poorer screening efficiency, as more non–relevant records needed to be screened to identify each relevant study. Based on WSS@100% values obtained for each dataset, Bos_2018 (A) and Schoot_2018 (C) were considered less challenging datasets, whereas Cohen_2006 (B) and Kwok_2020 (D) were considered challenging datasets. Each colored dot (blue or orange) represented a simulation. Three independent simulations were conducted for each ASReview LAB v.2 model (ELAS_u4 and ELAS_h3). Only the maximum mean value of intervening irrelevant articles, reflecting the lowest screening efficiency, was reported per model and per dataset, with corresponding standard deviations presented in parentheses. For each relevant article and dataset, the mean value across simulations was shown by the central line, while the upper and lower bounds depicted ±1 standard deviation.

The most advanced ELAS_h3 model showed that, in the *Kwok* dataset, low WSS@100% was primarily caused by three “hard-to-find” relevant articles, each requiring the screening of up to 254 irrelevant articles (Figure 1D). In contrast, in the *Cohen* dataset, a single “hard-to-find” relevant article required reviewers to examine 967 irrelevant articles (Figure 1B). In less challenging datasets, less than 100 irrelevant articles needed to be screened to find the “hardest-to-find” relevant article with ELAS_h3: 97 for *Bos* (SD 3) and 75 (SD 1) for *Van De Schoot* (Figures 1A and 1C). ELAS_u4 required more: 241 for *Bos* (SD 2) and 761 (SD 2) for *Van De Schoot*. Overall, ELAS_u4 and ELAS_h3 performed well in retrieving relevant articles early in the screening process, but even the most advanced ELAS_h3 model struggled with detection of “hard-to-find” article, requiring up to 967 irrelevant articles to be screened, more than nine times the number needed in less challenging datasets.

### Development of an EdRoSi AI Enhanced Priority Screening Pipeline

Given the observed limitations, a new priority screening pipeline, entitled EdRoSi AI, was developed to enhance retrieval efficiency of all relevant articles in challenging datasets. The hypothesis was that a small number of relevant and irrelevant articles, previously identified using static text-representation active learning methods, would be sufficient for effective fine-tuning of a biomedical transformer model to improve retrieval of relevant articles in challenging datasets (Figure 2).

**Figure 2:**
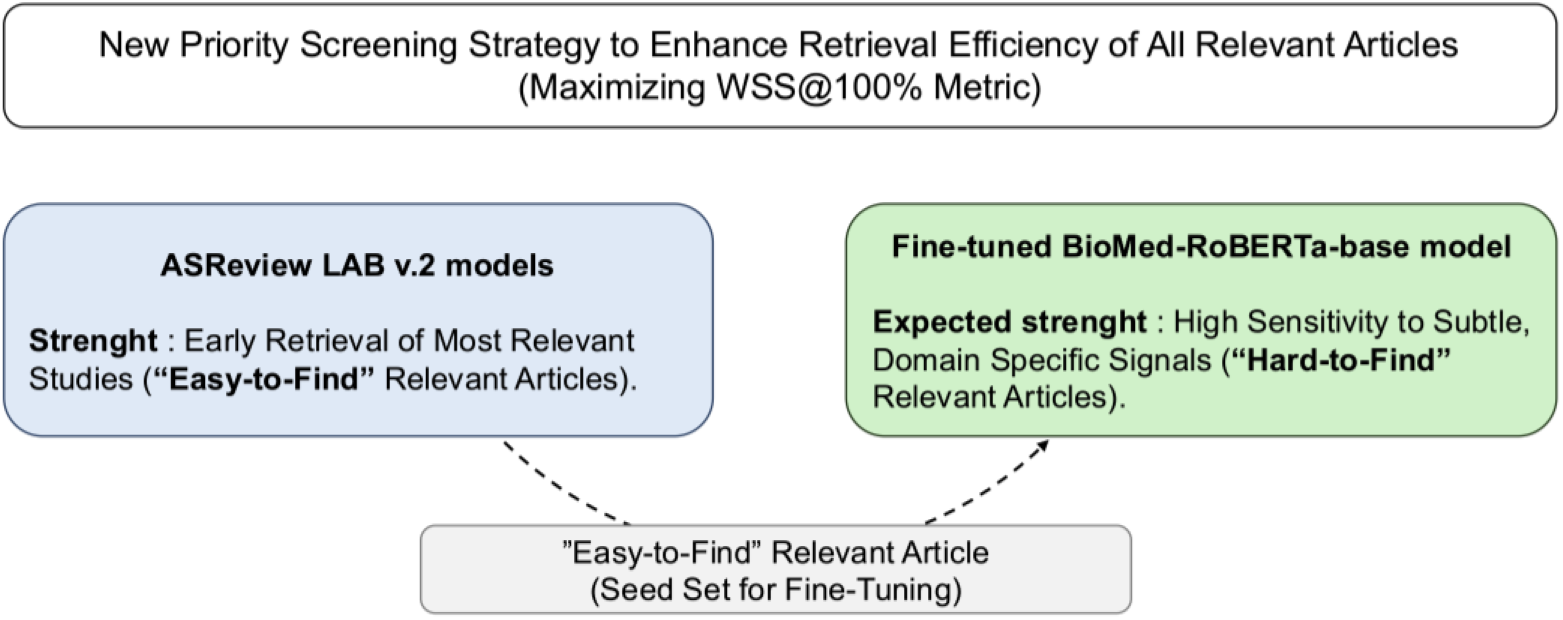
Development of EdRoSi AI, an Enhanced Priority Screening Pipeline. This schematic illustrated a hybrid priority screening approach that was designed to improve the retrieval of all relevant studies in a systematic review and was evaluated in this work. The strategy combined ASReview LAB v.2 models, which were particularly effective for the early identification of highly relevant and “easy-to-find” articles, with a fine-tuned BioMed-RoBERTa-base model that was optimized to detect subtle, domain-specific signals characteristic of “hard-to-find” relevant articles. The fine-tuning of the BioMed-RoBERTa-base model was performed using an initial set of “easy-to-find” relevant articles identified during early screening.

We showed in our study that ASReview LAB v.2 models (ELAS_u4 and ELAS_h3) performed well for early retrieval of most relevant studies (“easy-to-find” relevant articles) but showed limitations in identifying “hard–to–find” articles (Supplemental Table 1, Figures 1B and 1D). Both approaches rely on static text-representations, TF–IDF features for ELAS_u4 and transformer–based embeddings for ELAS_h3, both paired with SVM classifiers. Because these representations are generated independently of the screening task, they capture only generic semantic patterns and may miss subtle, domain–specific signals required to optimize stringent performance metrics such as WSS@100%.

### Efficiency Assessment of a Hybrid Priority Screening Approach Combining ASReview LAB v.2 Models Individually with a Fine-Tuned BioMed-RoBERTa-Base Model

To test this hypothesis, the BioMed-RoBERTa-base model was selected^25^. This language model, built upon the RoBERTa-base architecture, underwent continued pretraining on 2.68 million full-text biomedical papers from the Semantic Scholar corpus^26^. Its domain-specific design makes it particularly suitable for biomedical natural language processing (NLP) applications, including classification tasks.

The analysis focused on the *Cohen* dataset, which includes an exceptionally challenging relevant article that appears after screening 967 irrelevant records. This single “hardest–to–find” article substantially reduced the WSS@100% score, making it the primary target for methodological improvement. Our objective was to accelerate the detection of this article, and thereby increase WSS@100%, by fine–tuning BioMed-RoBERTa-base using relevant records efficiently identified by ELAS_h3.

Since the most challenging relevant article to locate was also the final relevant article retrieved, we hypothesized that fine-tuning the model using only the last 10 identified relevant articles, along with an equal number of hard irrelevant articles found among them, would optimize early-retrieval performance and yield both higher and more consistent WSS@100% results. When combined with hard irrelevant articles, this targeted fine–tuning is expected to sharpen discrimination precisely where current models tend to fail, enabling earlier attainment of full recall. The control conditions involved fine-tuning BioMed-RoBERTa-base using the last 10 relevant articles alongside randomly chosen irrelevant articles from the dataset. Additionally, it was fine-tuned with all 40 relevant articles, paired either with randomly selected irrelevant articles from the entire dataset or with hard irrelevant records.

WSS@100% improved substantially when BioMed-RoBERTa-base was fine–tuned, whether using all relevant articles or only the last 10 previously identified by ELAS_h3, and whether paired with randomly selected irrelevant articles or hard irrelevant articles, compared with using ELAS models alone. ELAS_h3 alone reached a WSS@100% of 46.5% (SD 0.0%), and ELAS_u4 reached 31.1% (SD 0.0%) (Figure 3). Fine–tuning with the 10 most recently identified relevant articles together with an equal number of hard irrelevant articles located between them produced the highest performance, achieving a WSS@100% of 83.3% (SD 0.4%). This slightly outperformed the configuration using all relevant articles paired with hard irrelevant studies (81.6%, SD 0.1%). In contrast, using randomly selected irrelevant articles resulted in lower and less stable performance: (i) Last 10 relevant + random irrelevant articles: 74.3% (SD 14.2%); (ii) All relevant (n = 40) + random irrelevant articles: 65.5% (SD 6.5%) (Figure 3).

**Figure 3.**
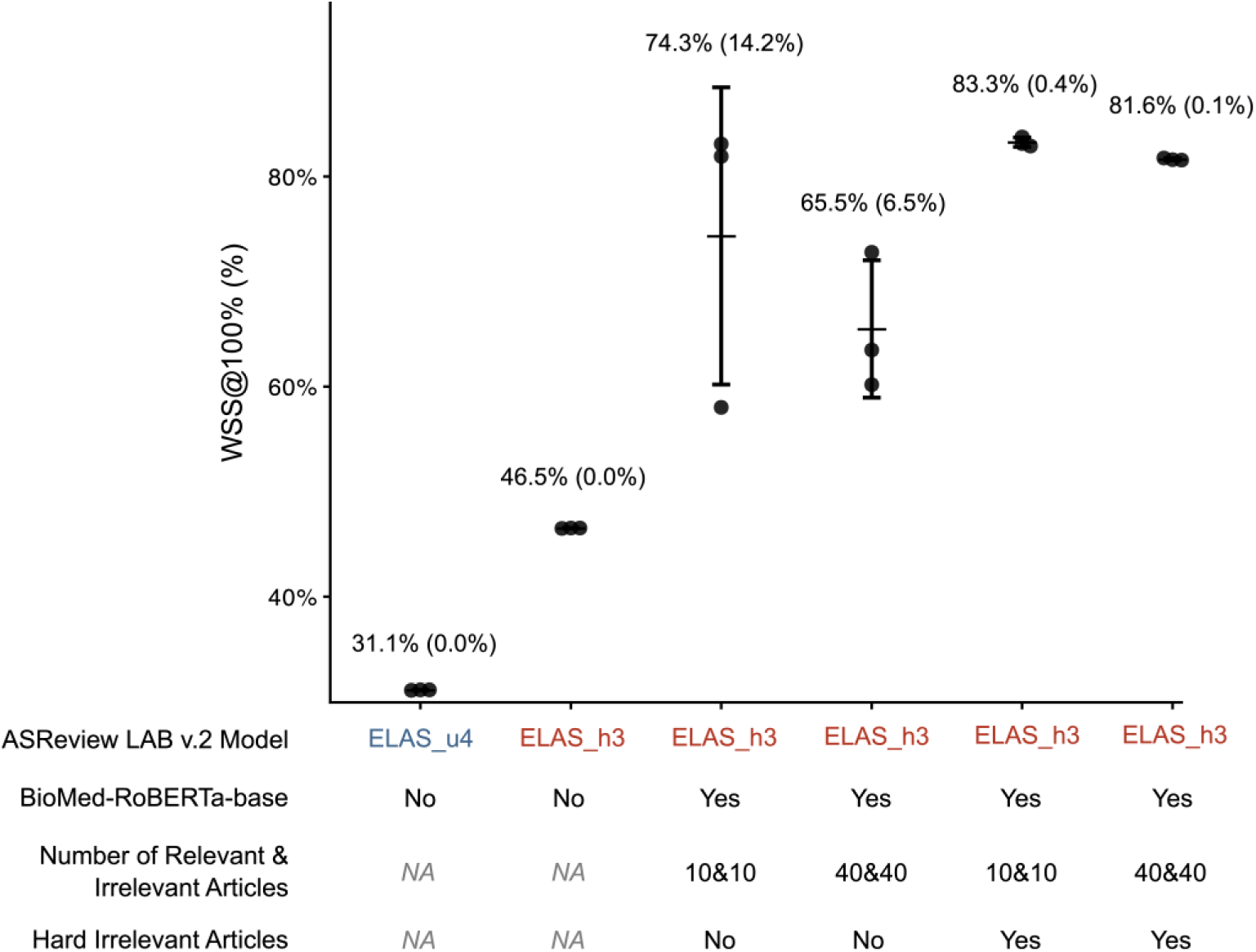
Efficiency Assessment of a Hybrid Priority Screening Approach Combining ASReview LAB v.2 Models Individually with a Fine-Tuned BioMed-RoBERTa-Base Model. The performance of the hybrid priority screening approach was evaluated using Work Saved over Sampling at 100% recall (WSS@100%), based on three independent simulations per condition. Four parameters were examined: (i) the type of ASReview LAB v.2 model used for early retrieval of relevant articles (ELAS_u4 or ELAS_h3); (ii) the use (Yes or No) of a fine-tuned BioMed-RoBERTa-base model; (iii) the number of relevant articles and a matching number of irrelevant articles used for fine-tuning (10 relevant articles and 10 irrelevant articles or 40 relevant articles and 40 irrelevant articles); and (iv) the inclusion (Yes or No) of hard irrelevant articles in the fine-tuning set, defined as irrelevant articles located between relevant articles. “NA” indicated “non applicable” because the model was not used. In this setting, WSS@100% corresponded to the articles screened during the ASReview ELAS_h3 workflow plus the additional articles screened following re-ranking by the fine-tuned BioMed-RoBERTa-base model. Each dot represented one simulation; mean WSS@100% values written above dots were depicted by the central line, while the upper and lower bounds depicted ±1 standard deviation (also written in parathesis after mean WSS@100% value).

With the optimal setup (last 10 relevant + hard irrelevant articles), just 2.1% of unscreened records needed screening (Supplemental Table 2). This improvement came at minimal computational cost: fine–tuning and inference with BioMed-RoBERTa-base required an average of 22 minutes, in addition to 54 minutes for ELAS_h3, when run on a MacBook Pro 2020 (2 GHz Quad–Core Intel Core i5, 16 GB RAM).

Fine-tuning BioMed-RoBERTa-base with the last 10 relevant articles and hard irrelevant study titles and abstracts greatly boosted WSS@100% over ELAS models, without much extra computation. Using hard irrelevant articles had the most impact on detecting elusive articles, and limiting fine-tuning to recent relevant articles led to modest gains in retrieval performance.

### Proposed EdRoSi AI Priority Screening Pipeline for Enhanced Title and Abstract Screening

Based on these results, EdRoSi AI, a new priority screening pipeline is proposed for situations where maximum recall is required (Figure 4):

1. Employ the ELAS_h3 model from ASReview LAB v.2 that showed superior WSS@100% on both challenging and less challenging datasets, compared to ELAS_u4 model.
2. Articles should be screened, with the process discontinued once 100 consecutive irrelevant articles have been identified. The rationale is based on findings that fewer than 100 irrelevant articles were required to locate the “hardest-to-find” article in non-challenging datasets when using ELAS_h3.
3. If at least 10 relevant articles have been identified efficiently with ELAS_h3, proceed to fine-tune BioMed-RoBERTa-base using the last 10 relevant articles and the same number of hard irrelevant articles located between them.
4. Refine the screening and data-driven stopping rule; screening 2.1% of new articles could serve as a starting point. If less

**Figure 4.**
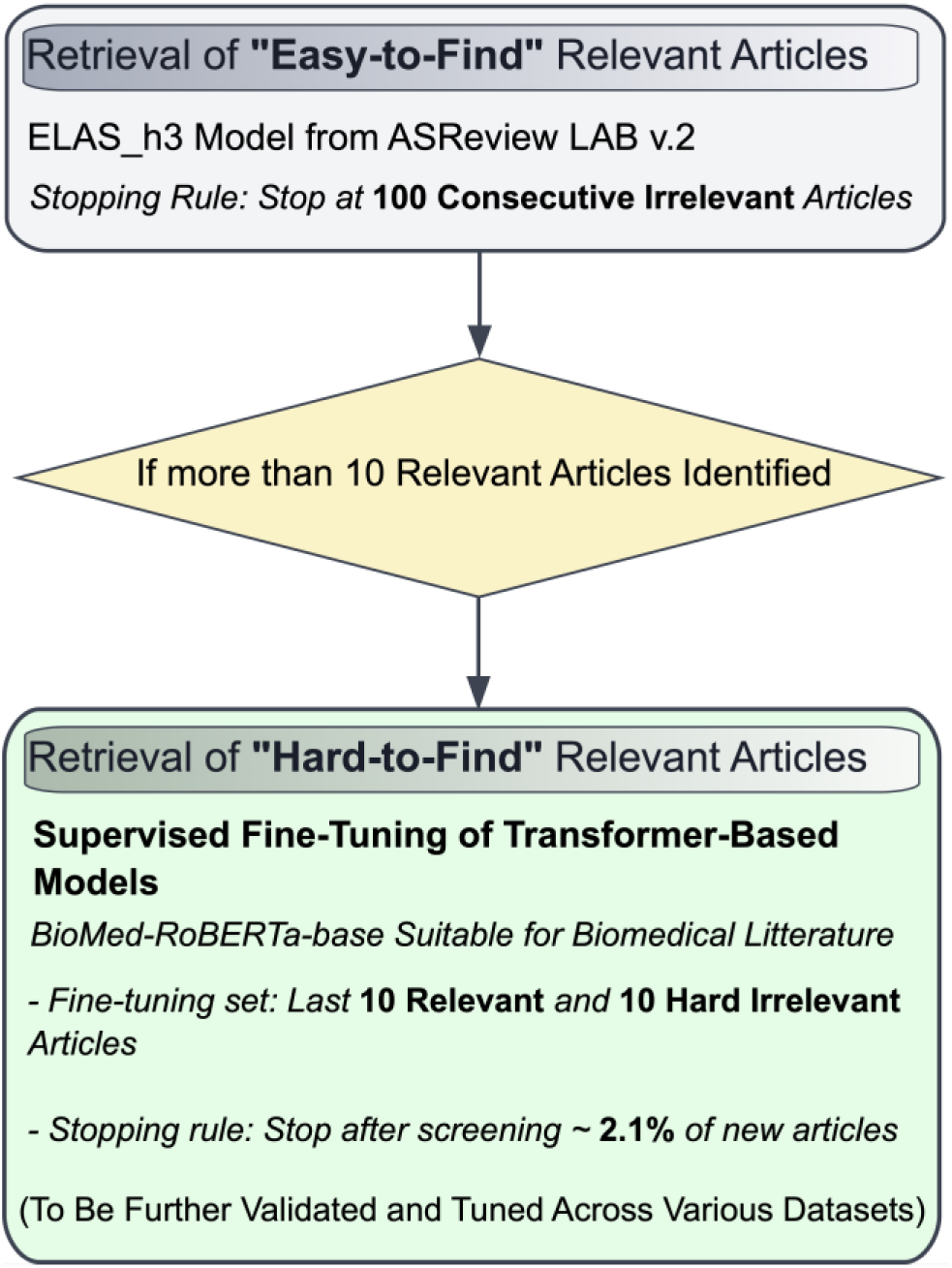
Proposed EdRoSi AI Priority Screening Pipeline for Enhanced Title and Abstract Screening. This schematic illustrates the EdRoSi AI priority screening pipeline designed to improve retrieval efficiency in systematic reviews. The workflow begins with the retrieval of “easy–to-find” relevant articles using the ELAS_h3 model from ASReview LAB v.2, with screening stopped after 100 consecutive irrelevant articles. If more than 10 relevant articles are identified, the pipeline transitions to a second stage targeting “hard–to–find” relevant articles through supervised fine–tuning of a transformer–based model (BioMed–RoBERTa–base), which is suitable for biomedical literature. Fine–tuning is performed using the most recent 10 relevant articles and 10 hard irrelevant articles, defined as irrelevant articles located between relevant articles in the ranking. Screening in this stage stops after approximately 2.1% of new articles (not yet screened). Such new pipeline must be validated and tuned across multiple datasets.

## Discussion

The results of this study show that while ASReview LAB v.2 models, particularly the transformer–based ELAS_h3, performed well for early retrieval of most relevant studies, they continue to struggle with detecting a small number of “hard–to–find” articles that dominate the tail of the ranking and strongly depress WSS@100%. Supervised fine-tuning of transformer-based models pre-trained on biomedical literature is a promising approach^6,27,28^. It allows adaptation of the model’s internal representations to the specific inclusion criteria of a review. That potentially improves sensitivity to subtle relevance signals and enhancing detection of “hard-to-find” articles^27^. We showed in this study that supervised fine–tuning of BioMed–RoBERTa–base with relevant articles efficiently identified with ELAS_h3, substantially improved the detection of these elusive articles. This highlights the added value of transformer–based classification models, which adapt their internal representations to subtle, domain–specific relevance patterns that static text embeddings cannot capture.

Using all available relevant records may bias the model toward dominant semantic patterns, whereas focusing on the most recently found relevant articles exposes it to rare, heterogeneous, and borderline cases. Indeed, surprisingly, just 10 relevant late-stage articles and 10 hard irrelevant articles were sufficient to fine-tune a transformer-based classifier, greatly improving retrieval of “hard-to-find” articles, far fewer than the thousand usually needed at lowest for effective transformer fine-tuning^6,27,28^. This unexpected efficiency may be explained by the unusually high information density of the last relevant articles, which are precisely those representing rare, heterogeneous relevance signals. It can also be explained by the discriminative power of hard irrelevant articles drawn from ambiguous semantic neighborhoods. It would be interesting for future studies to investigate the minimal number of relevant and irrelevant articles required to achieve effective retrieval of “hard-to-find” articles.

Given the strong performance achieved with targeted fine–tuning, one might consider iterative fine–tuning of RoBERTa earlier in the screening process; however, such an approach would likely incur substantial computational cost at every iteration, and its marginal benefit over static active–learning models remains uncertain. For these reasons, our findings suggest that fine–tuning a domain–specific transformer should primarily be used as a final–phase strategy to accelerate retrieval of the remaining hard–to–find articles once a minimal training signal is available. Such EdRoSi AI priority screening pipeline could be particularly valuable for situations where maximum recall is required. It could be particularly useful in scoping reviews addressing broad or heterogeneous topics, where relevance signals are inherently subtle and diffuse, and may not be adequately captured by traditional word-frequency–based or generic embedding-based approaches.

Future research should focus on developing and validating a data-driven stopping criterion for concluding screening following RoBERTa fine-tuning. Moreover, it is important to acknowledge that a major limitation of this study lies in the evaluation of the hybrid priority screening strategy on a single challenging dataset. To establish its generalizability, further testing across multiple complex benchmark datasets exhibiting diverse semantic irregularities and tail-risk behaviors is necessary.

## Supplementary Tables

**Supplemental Table 1:** Performance of ASReview LAB v.2 ELAS_h3 and ELAS_u4 Models on Challenging and Less challenging Datasets. Work Saved over Sampling at 100% recall (WSS@100%) obtained across three independent randomizations for ASReview LAB v.2 ELAS_h3 and ELAS_u4 models, with mean WSS@100% values and standard deviation reported for each challenging and less challenging validation dataset.

| Validation dataset | Screening tools | Randomization | WSS@100% (%) | Mean WSS@100% | Standard deviation WSS@100% |
| --- | --- | --- | --- | --- | --- |
| Bos_2018 | ELAS_u4 | Randomization 1 | 89.57320367 | 89.53718711 | 0.062382525 |
|  |  | Randomization 2 | 89.57320367 |  |  |
|  |  | Randomization 3 | 89.46515397 |  |  |
|  | ELAS_h3 | Randomization 1 | 94.59751486 | 94.59751486 | 0.036016568 |
|  |  | Randomization 2 | 94.63353142 |  |  |
|  |  | Randomization 3 | 94.56149829 |  |  |
| Van_De_Schoot_2018 | ELAS_u4 | Randomization 1 | 83.0335524 | 83.05084746 | 0.017295054 |
|  |  | Randomization 2 | 83.06814251 |  |  |
|  |  | Randomization 3 | 83.05084746 |  |  |
|  | ELAS_h3 | Randomization 1 | 95.03631961 | 95.10549983 | 0.135078687 |
|  |  | Randomization 2 | 95.26115531 |  |  |
|  |  | Randomization 3 | 95.01902456 |  |  |
| Cohen_2006 | ELAS_u4 | Randomization 1 | 31.0927673 | 31.11897275 | 0.022694586 |
|  |  | Randomization 2 | 31.13207547 |  |  |
|  |  | Randomization 3 | 31.13207547 |  |  |
|  | ELAS_h3 | Randomization 1 | 46.50157233 | 46.52777778 | 0.022694586 |
|  |  | Randomization 2 | 46.5408805 |  |  |
|  |  | Randomization 3 | 46.5408805 |  |  |
| Kwok_2020 | ELAS_u4 | Randomization 1 | 30.95525998 | 30.98213086 | 0.023270869 |
|  |  | Randomization 2 | 30.9955663 |  |  |
|  |  | Randomization 3 | 30.9955663 |  |  |
|  | ELAS_h3 | Randomization 1 | 45.14308746 | 45.08934569 | 0.046541739 |
|  |  | Randomization 2 | 45.06247481 |  |  |
|  |  | Randomization 3 | 45.06247481 |  |  |

**Supplemental Table 2:**
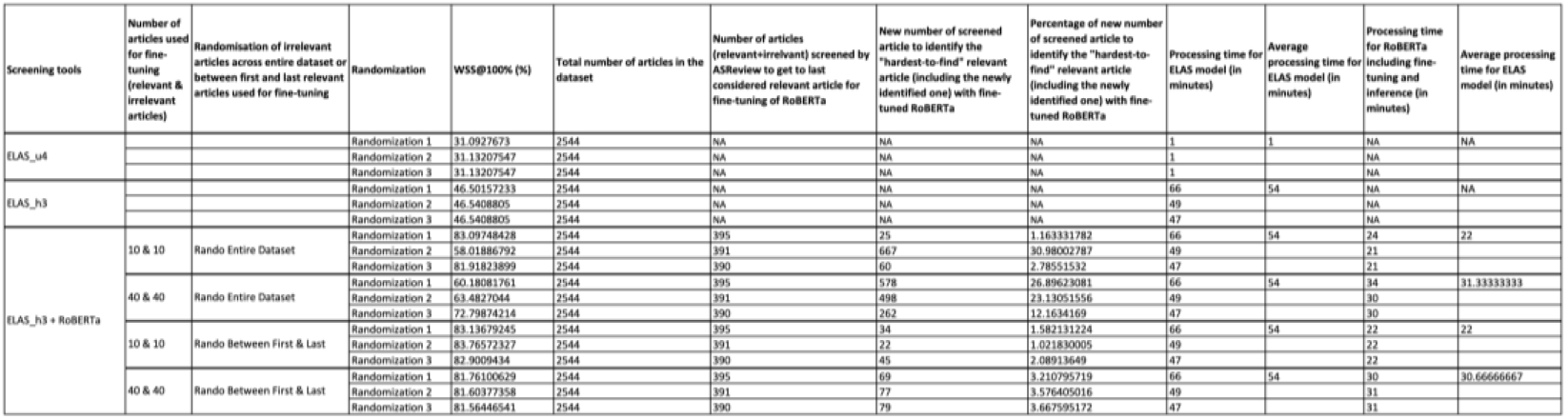
WSS@100% and Processing Time for ASReview LAB v.2 Alone or In Combination with EdRoSi AI for *Cohen* Challenging Dataset. Work Saved over Sampling at 100% recall (WSS@100%) and processing times obtained for the *Cohen* challenging dataset using ASReview LAB v.2 models alone (ELAS_u4 or ELAS_h3) or in combination with the EdRoSi AI hybrid priority screening approach, across three independent randomizations and multiple fine-tuning configurations.

## Data Availability

All data produced in the present study are available upon reasonable request to the authors

## Acknowledgments

This work was supported by Boiron.

## Declaration of generative AI and AI-assisted technologies in the manuscript preparation process

During the preparation of this work the authors used Copilot in order to improve grammar and sentence structures. After using this tool/service, the authors reviewed and edited the content as needed and take full responsibility for the content of the published article.

